# DBScope: a versatile computational toolbox for the visualization and analysis of sensing data from Deep Brain Stimulation

**DOI:** 10.1101/2023.07.23.23292136

**Authors:** Andreia M. Oliveira, Eduardo Carvalho, Beatriz Barros, Carolina Soares, Manuel Ferreira-Pinto, Rui Vaz, Paulo Aguiar

**Affiliations:** Neuroengineering and Computational Neuroscience Lab, Instituto de Investigação e Inovação em Saúde (i3S) - University of Porto, Portugal; Faculty of Engineering of University of Porto (FEUP), Portugal; ICBAS – School of Medicine and Biomedical Sciences – University of Porto, Portugal; Faculty of Medicine of University of Porto (FMUP), Portugal; Centro Hospitalar Universitário de São João (CHUSJ), Portugal

**Keywords:** Deep Brain Stimulation, Data-driven therapies, Electrophysiological biomarkers, Computational tools, local field potentials, Percept PC

## Abstract

Deep brain stimulation (DBS) is a therapy applied in numerous neurologic diseases, yielding major improvements in quality of life. Current implantable devices can record brain activity (in the form of local field potentials) at the site of stimulation, providing a window into the neuropathological phenomena and the potential to improve clinical care. Data-driven therapies often rely on tools to process, visualize, and analyze the data. However, existing tools in DBS are yet to fully exploit the devices’ sensing capabilities. We aimed to develop a user-friendly open-source toolbox for the visualization and analysis of sensing data from DBS. Special attention was given to enrich this toolbox with features which would foster its usefulness in both clinical and research environments. We developed a standalone MATLAB-based toolbox, called DBScope, capable of parsing the data generated by current sensing devices and producing rich visualizations with clinical and research relevance. The toolbox can be operated in two ways: through a user interface, bypassing programming experience requirements; and, programmatically, acting as a library of functions, which can be further adapted to user needs. We provide a detailed description of the toolbox features and exemplify its application in two case studies. DBScope is an open-source toolbox that provides visualization and analytical tools for clinical and research use, paving the way towards the improvement of data-driven DBS therapies. Additional functionalities are being considered for future updates.

## Introduction

Deep Brain Stimulation is an established practice for the treatment of neurologic diseases and shows promising prospects for psychiatric conditions.^1, 2^ It involves the delivery of electrical stimulation to targeted brain regions, that vary in respect to the disease in question.^3-5^ While the exact mechanisms of action underlying DBS remain unknown, evidence supports its capacity to modulate neural circuits locally and non-locally.^5, 6^ Current DBS therapies are delivered in a continuous and open-loop fashion: using a trial and error approach, the clinician selects the best performing stimulation parameters (i.e., a combination of higher symptom mitigation with lower incidence of adverse effects) from established therapeutic parameter ranges;^7-9^ after selection, the parameters are kept until the following clinical session, where they are re-adjusted.

Medtronic, a leading company in DBS, announced in 2020 the first US/EU approved neurostimulator with the ability to chronically sense local field potentials during stimulation (Percept PC).^10^ Since then, clinicians and researchers have included the retrieval of sensing data (using a ‘programming’ tablet) on the standard-of-care. This inclusion has been key in increasing our understanding of disease-related brain activity patterns, their temporal evolution, and their modulation in response to therapies. In addition, it also gave the opportunity to unveil new electrophysiological biomarkers and ultimately bring the implementation of adaptive stimulation therapies closer to clinical practice.^11-13^ Nonetheless, the current visualization tools accessible on the clinician’s tablet are limited: they fall short in fully exploiting the information embedded within the recordings and are ill-suited for longitudinal analyses (reviewing current and previous data). Consequently, clinicians and researchers have been compelled to seek and develop custom-made software to handle the files exported from the tablet.

Until recently, very few toolboxes were created for this purpose and addressing only simple extraction and visual methods for specific types of recordings.^13, 14^ Yet, their use requires expertise in programming, a deterrent for many DBS experts. This requirement has recently been challenged by a new open-source platform called BRAVO (Brain Recording Analysis and Visualization Online), which using a web-based interface offers visual tools for DBS data analysis.^15^ While this platform addresses the ‘parsing challenge’ and allows the concurrent analysis of several files, its use poses technical challenges (requires confi.guration of a Linux server), and special attention regarding clinical data sharing/security (non-local data analysis).

In short, there is still a pressing need for easy-to-use yet versatile tools that facilitate the visualization and analysis of the data downloaded from currently available neurostimulators. To answer this need, we developed a free open-source toolbox, named DBScope, that imports data from neurostimulation devices and can be operated in two ways: via user interface and programmatically, as a library of functions. In this way, it can be used by both clinicians during clinical sessions (for instance, to visually inspect data from the current or previous in-clinic visits), and by researchers in their research pipelines (e.g., for pre-processing, feature extraction and biomarker search). At this stage, DBScope contains a parser for the Percept PC device, but as other devices with sensing capabilities become available, other data parsers can be included. All in all, the DBScope toolbox is set to facilitate the clinical decision-making process and the identification of clinically relevant biomarkers.

## Methods and Results

### DBScope conception and core features

DBScope was developed with MATLAB 2022b (MathWorks, Natick, MA), a well-established scientific programming language with several built-in tools, ranging from signal analysis to user interface creation. The initial motivation for the toolbox stemmed from an interest in conducting in-depth analysis on the files extracted from the Medtronic’s Percept PC setup. Besides information regarding the in-clinic visit, the patient and the neurostimulator, these files contain two main groups of recordings: the in-clinic recordings, which are subdivided in *Survey, Setup*, and *Streaming* sensing modes; and the chronic (or out-of-clinic) recordings, containing the *Timeline* and the *Events* sensing modes (up to 60 days for the Percept PC device).

The DBScope toolbox was designed with a set of core features in mind: 1) interactivity, as to ensure that users with little to no programming experience could use it in their practices; 2) reusability, so that users with some programming experience could easily create or adapt the tools to better fit their research questions; 3) integration, to allow the simultaneous analysis of both different signals (in cases where motor data is also acquired) and multiple files (for longitudinal studies); 4) customization, to the extent that each sensing mode should have tailored methods. We used an object-oriented programming paradigm to facilitate the fulfillment of these requirements (Figure 1). Specific functions in DBScope were derived from pre-existing open-source computational toolboxes: the extraction of LFP and stimulation information methods are based on the analogous functions present in the Perceive toolbox;^13^ the electrocardiographic (ECG) artifact cleaning tool was extracted from the Percept toolbox.^14^

**Figure 1.**
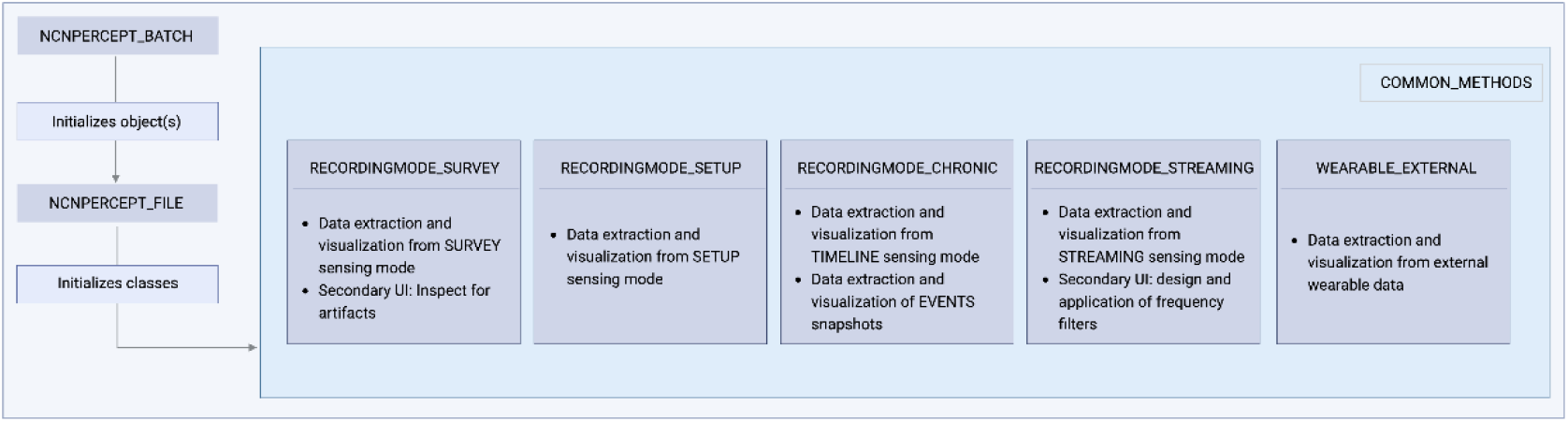
*DBScope* internal structure. The toolbox was designed using an object-oriented programming framework. Upon initialization, the class NCNPERCEPT_BATCH separates the loaded files in patients (each containing one or several files). Each file gives rise to a NCNPERCEPT_FILE object. These objects contain six classes, each with its own properties and methods. Each class is named after the type of recording that it can access. The WEARABLES_EXTERNAL class handles additional data that is not present within the neurostimulator files and needs to be loaded separately afterward. COMMON_METHODS is an auxiliary class that runs background methods.

### User interface

Integrating information from multiple types of recordings can be troublesome and time-consuming. To facilitate this process, we developed a user interface (Figure 2), which provides a graphical environment for data analysis and exploration. This inclusion is mainly addressed to an audience with no previous experience in programming, that is keen on including DBS data analysis into their clinical or research routines. Nevertheless, the toolbox also enables experienced programmers to create their own scripts, adapting or creating new functions to better fit their research questions (Supplementary Figure S1).

**Figure 2.**
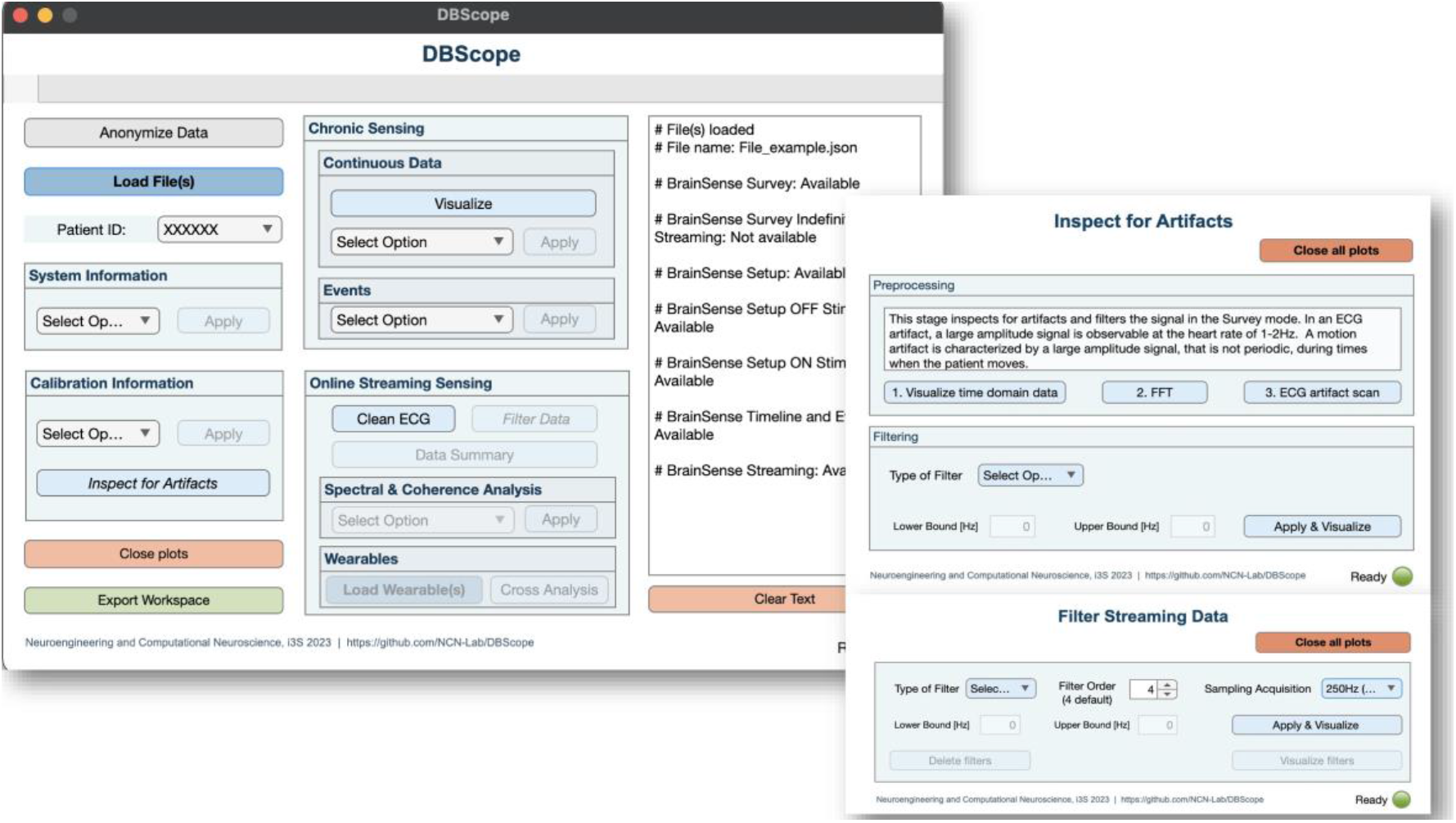
DBScope graphical interface. The toolbox comprises a main interface that accommodates four sections. Each section gives access to part of the data contained in the loaded files. Within the *Calibration Information* and *Online Streaming Sensing* sections, the user can access secondary interfaces that are used in artifact inspection and signal filtering, respectively. The main interface also contains a text area that displays information during user interaction. For instance, upon loading a file, it shows the available sensing modes.

### Operational Workflow

The first step in DBScope is to load the files of interest. The toolbox is capable of handling one or multiple files from the same patient, facilitating combined/aggregated analysis as required. Once the files are loaded, four main streams of operations are suggested (Figure 3), considering the sensing modes present in the loaded file(s):

- **System Information** provides information on the neurostimulator, the patient, and the in-clinic visit.
- **Calibration Information** gives access to the impedance tests, to the *Survey* and *Setup* recordings, and to a secondary interface for artifact exploration.
- **Chronic Sensing** offers visualization and analytical tools for the *Timeline* and *Events* recordings.
- **Online Streaming Sensing** draws on four main functionalities: 1) an ECG cleaning algorithm; 2) a filtering tool that calls a secondary interface; 3) spectral and correlation tools; and 4) a tool for visual cross-analysis with wearable data. For the latter, the user must load the corresponding wearable data files, in a CSV format (for more information, consult the DBScope documentation). During visualization, the toolbox automatically aligns the wearable recordings with the streaming recordings using the marked timestamps.

**Figure 3.**
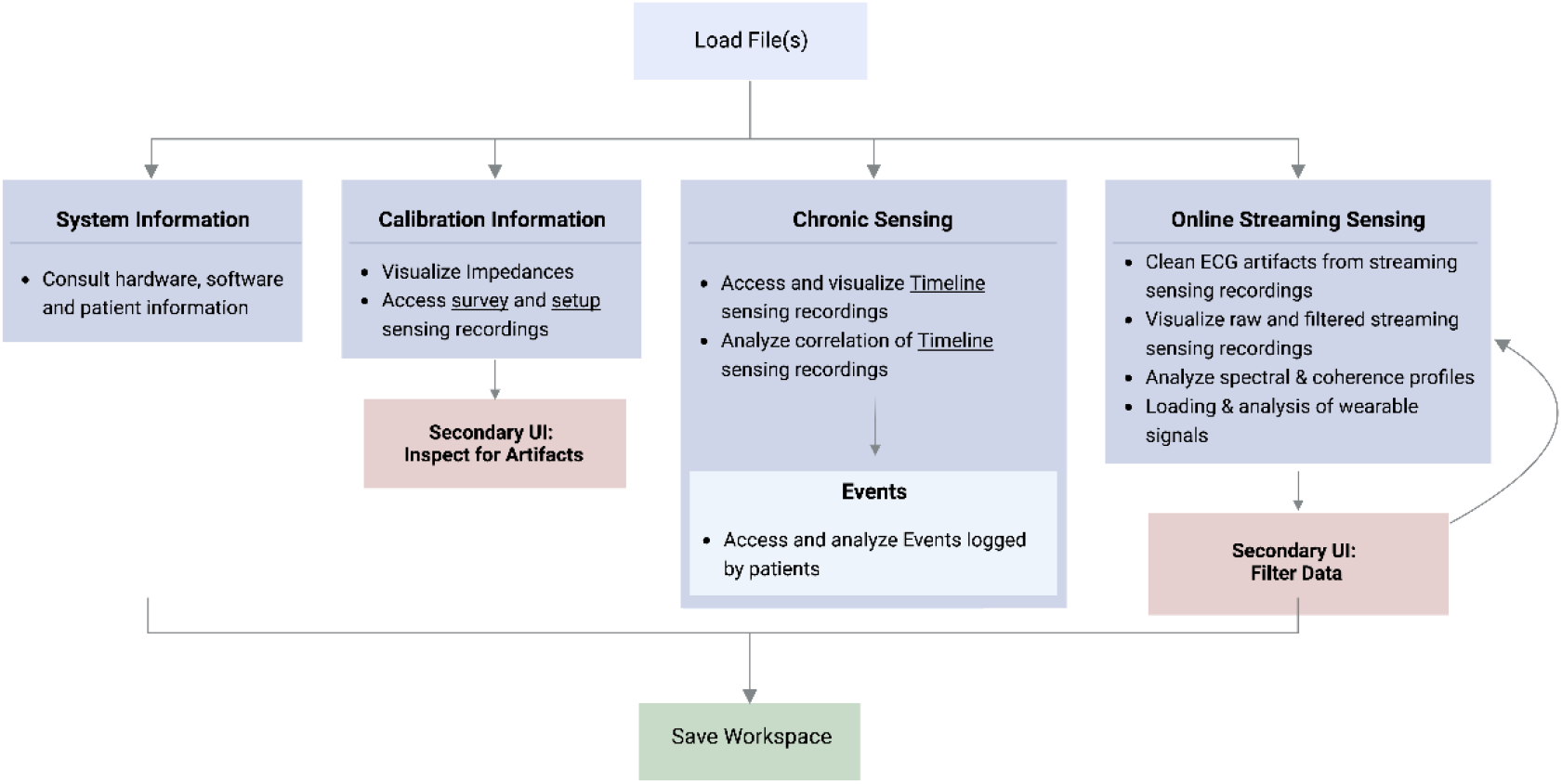
General *workflow*. The user starts by loading one or multiple files. The user can always return to this step, with the option to save the current workspace. After loading the file(s), four streams of operations are available: **System information**, gives information about the patient, the device, and the in-clinic visit; **Calibration Information** deals with the recordings obtained when testing and setting the stimulation and sensing parameters, and has a secondary interface for artifact inspection; **Chronic Sensing** offers multiple visual and analytical functions for the Timeline and Events recordings; **Online Streaming Sensing** provides tools for visualization, artifact removal, filtering (using a secondary interface), and alignment with wearable data. Regardless of the chosen workflow, the user can always save the workspace (for more information, consult the DBScope documentation).

**Figure 4.**
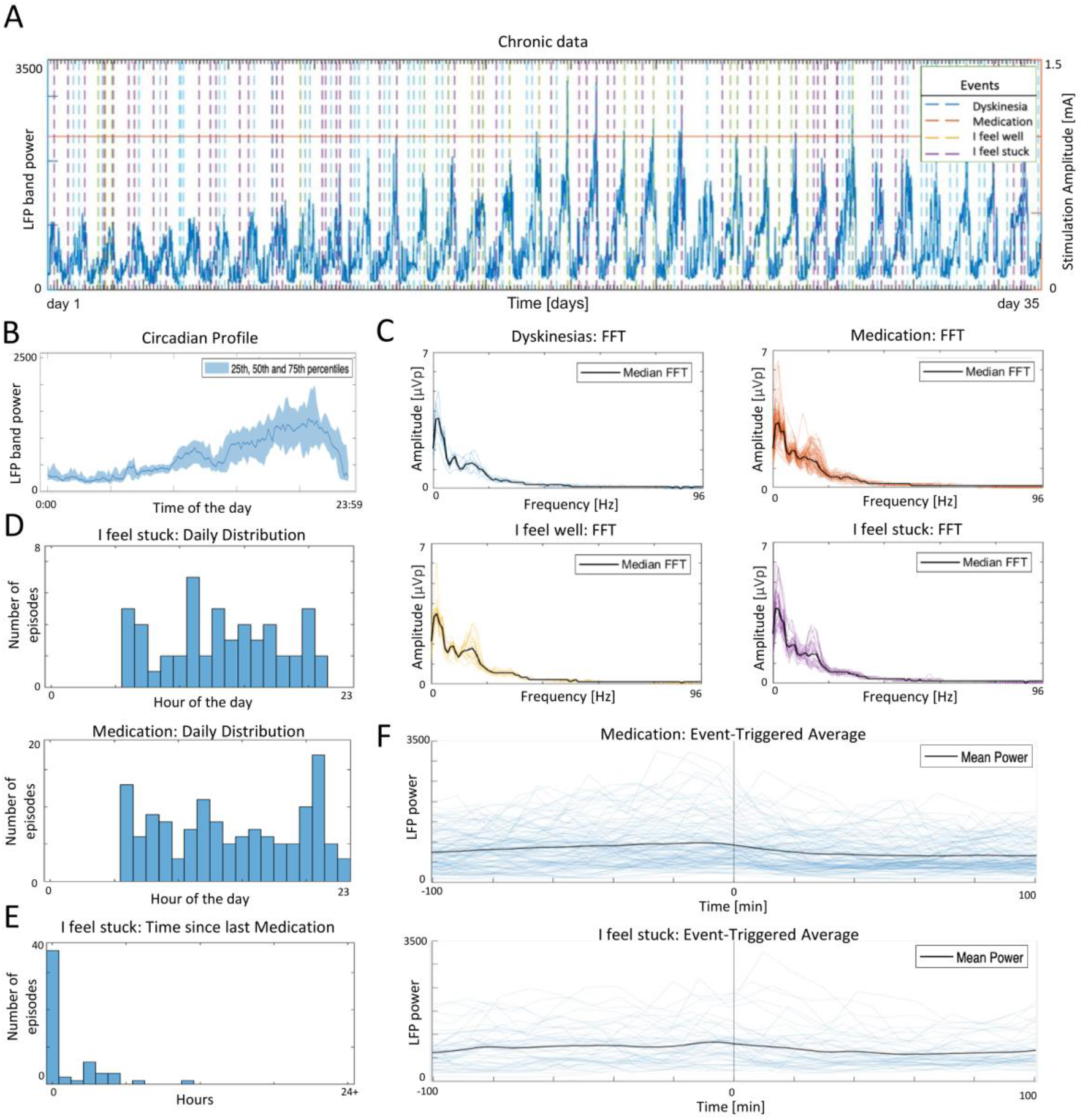
Case study of out-of-clinic data. **(A)** Representation of *Timeline* and *Events* of the selected hemisphere. The recordings cover approximately one month in a conventional DBS protocol (continuous fixed stimulation amplitude). **(B)** Estimated circadian profile of the *Timeline* recording in (A). The LFP power is higher during waking hours. **(C)** Fast Fourier Transform (FFT) profiles for each of the recorded *Events*. The observed median profiles were similar, with the highest variation around the 15 Hz peak. **(D)** Daily distribution of ‘Medication’ and ‘I feel stuck’ episodes. Notice that distributions are similar, with three periods of higher incidence (6h, 11-12h, 20-21h). **(E)** Distribution of the time between consecutive ‘Medication’/’I feel stuck’ pairs. Interestingly, ‘I feel stuck’ episodes are mostly reported after ‘Medication’. **(F)** Event-triggered average plots showing the power of the selected band in the proximity of ‘Medication’ and ‘I feel stuck’ episodes. The mean power decreases after the two events.

### Case-Studies

To exemplify the application of the toolbox and showcase some of its features (for more, see Supplementary Figure S2), we present two case studies: the first about the long-term LFP dynamics (up to a month) of out-of-clinic recordings; the second about the LFP dynamics in shorter time scales (in the domain of minutes) of in-clinic recordings.

### Case study: out-of-clinic recordings

This case study investigates a single hemisphere of one patient. Circadian and diurnal power fluctuations within the preselected frequency band can be observed throughout the entire 34-day *Timeline* recording (Figure 5A-B). While beta power exhibits a consistent reduced magnitude overnight (which is indicative of sleep time), most fluctuations occur during daytime and increase in range over the recording period. The latter phenomenon may be explained by the fact that the implantation took place two weeks prior to the recording and the tissue around the implant was still stabilizing (i.e., inflammatory response, edema, and scar tissue formation persisted).

**Figure 5.**
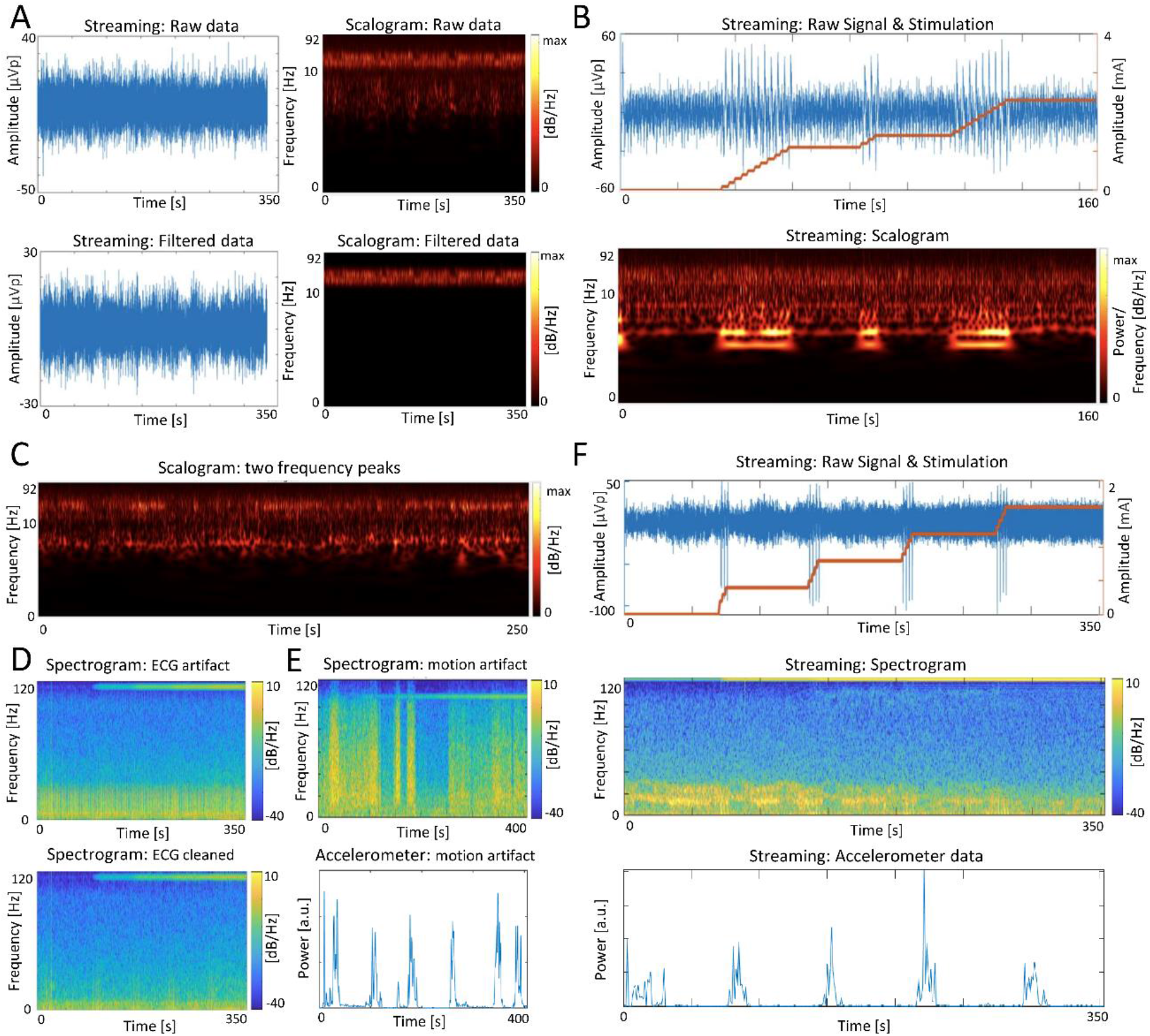
Case study of in-clinic recordings. **(A)** Application of a customized filter to better highlight a band of interest (13-35 Hz). **(B)** Raw neuronal recording and time-frequency analysis aligned to therapy changes. Amplitude stepping induces changes both at the raw signal (abrupt magnitude rises) and scalogram (high low-frequency activity). Note that the y-axis of the scalograms in (A) and (B) has a logarithmic scale. **(C)** Example of a recording where both beta and theta-alpha activities are elevated. **(D)** Recording with cardiac artifacts before (top) and after (bottom) cleaning. **(E)** Recording with motion-induced artifacts. The artifacts span across the entire frequency spectrum, being more intense at the peaks in motion power and vanishing once the motion power is plotted over zero. **(F)** Cross-signal analysis for a titration recording during a motor task. Both stimulation and movement suppress beta-band activity.

This patient’s *Events* recordings include ‘Medication’, ‘I feel well’, ‘I feel stuck’, and ‘Dyskinesias’ episodes (Figure 5C). Dyskinetic episodes are characterized by lower theta-alpha and beta powers (Supplementary Figure S3). The daily distribution of ‘I feel stuck’ episodes contains multiple periods of higher incidence, which are aligned with the medication intake distribution (Figure 5D). Interestingly, most ‘I feel stuck’ episodes appear immediately after medication (Figure 5E). While the inverse relationship would be expected (i.e., medication would decrease rigidity), it is possible that the patient was using the medication schedule as a cue to report the immediately preceding rigidity states.

Post-medication power shows a consistent decrease within a few minutes of medication intake (Figure 5F). This phenomenon aligns with the known suppression of beta activity induced by Parkinsonian medication and supports the usefulness of this frequency band in investigating the wear-off effect. This decrease is also seen after ‘I feel stuck’ episodes, which is not surprising given the temporal relationship with medication episodes (Figure 5E).

### Case study: in-clinic recordings

This case study explores the streaming recordings of multiple patients. Scalograms, obtained with a wavelet transform method, exhibit bands with heightened activity for many patients (Figure 5A, top plots). To enable easier analysis and interpretation aligned with clinical observations, a filtering tool can be applied to highlight and isolate these bands (Figure 5A, bottom plots). Instead of merely displaying the overall power of the selected band (like the ‘programming’ tablet), it exhibits distinct frequency variations within the band. Moreover, it provides greater flexibility by allowing the highlighting of multiple bands, which is advantageous in situations where monitoring a single band may not be sufficient for therapy evaluation (e.g., when the theta-alpha bands are also elevated, Figure 5C).

In many of the recordings, the stepping of stimulation amplitude resulted in abrupt rises of the LFP signal, evident by an increase in lower frequency oscillations (Figure 5B). This phenomenon has an artifactual nature and has been previously reported in Percept PC patients.^16^ Incidentally, artifacts in DBS are common and can arise from various sources, including cardiac activity (Figure 5D, top plot), motion (Figure 5E), and stimulation itself. Furthermore, it is not uncommon for a signal to be contaminated by multiple types of artifacts. In some recordings, the ECG cleaning tool can successfully remove the ECG artifact (Figure 5D, bottom plot).

In some patients, an increase in stimulation amplitude results in beta-band activity suppression both at rest (not shown) and during motor performance (Figure 5F). Additionally, desynchronization of beta activity is shown to occur at lower stimulation amplitudes during a motor task involving the opening and closing of the contralateral fist.

## Discussion

In this work, we address the need of accessible and comprehensive analytical tools for the DBS therapies. We present a toolbox with an intuitive user interface that allows both clinicians and researchers to access and explore intricate electrophysiological data. To the best of our knowledge, no other toolbox offers a comparable range of functions in terms of both extent and variety, together with ease to run/setup and customize (but see^15^). Among these functions, we would like to emphasize the inclusion of an ECG cleaning tool and the capability to visualize streaming recordings with data obtained from wearable devices. Furthermore, the toolbox operates without the need for an internet connection and patient data is not required to be stored on a server. While we acknowledge that a central database may be desirable in a healthcare setting, it is also true that it imposes additional technical/security measures regarding the sharing of personal data.

To standardize some of the most performed analyses of the field, we designed a novel structure of classes and developed a diverse repertoire of functions within each class. Some of these functions were adapted from existing toolboxes, notably in the detection and cleaning of cardiac artifacts. By adopting an object-oriented programming framework we were able to produce a toolbox that can analyze multiple files in a simple and intuitive workflow. This is desirable not only from a software design perspective, but also from a scientific perspective, where longitudinal, multi-signal and multi-patient analyses are relevant.^11, 17, 18^

To illustrate the application of DBScope, we present two case studies that concentrate on out-of-clinic and in-clinic recordings. The latter are more commonly used in the context of Percept PC, as they offer a higher temporal resolution. Nevertheless, out-of-clinic recordings still provide an invaluable glimpse into real-world settings, where data-driven DBS therapies aim to actuate. In fact, we observed that chronic data contains valuable information, such as the patients’ circadian patterns, which exhibit longer temporal dynamics. These patterns are challenging to identify in in-clinic recordings but are discernible in long-term ones. At the same time, it was possible to study the response of the LFP to specific occurrences, such as medication intake or rigidity episodes. While the marking of medication episodes showed some potential in monitoring the wear-off effect, other events were reported less times and typically coincided with medication intake, suggesting that the patient was reporting retroactively. Regarding the in-clinic recordings, the toolbox offered clear visualizations that facilitated the identification of both artifacts and clinically relevant frequency bands. Additionally, the possibility of aligning streaming recordings with wearable data proved to be a relevant addition to DBScope, enabling the investigation of movement-related modulations.

Although DBScope succeeded on many fronts, it bears some limitations. Presently, the toolbox operates exclusively on files extracted from Medtronic’s Percept PC setup. As new devices with sensing capabilities have since emerged (Medtronic’s Summit RC+S and Newronika’s AlphaDBS), we are considering the development of specific parsers to accommodate them. DBScope is also reliant on commercial software (MATLAB). However, it should be noted that universities and research institutes often offer campus-wide licenses to their members.

Future developments aim to address two challenges. The first lies in the search for biomarkers. Defining a biomarker, or library of biomarkers, is an essential, albeit complex, step for the development of patient- and symptom-specific DBS therapies.^19-21^ A direct mapping between brain activity and reported symptoms is seldom possible, due to the subjective and elusive natures of the latter. In these cases, a second signal, more interpretable and highly linked with the symptom type, is often introduced to facilitate the mapping process. For instance, accelerometry data is widely used to link electrophysiological signals with motor symptoms. In light of this, we have already included the functionality to load accelerometry data into DBScope and are actively developing additional methods to leverage the information contained in these signals. The second challenge stems from the fact that LFP are prone to artifacts of different origins, such as cardiac, movement, and stimulation. Although current devices have implemented artifact cleaning algorithms, not all artifacts can be reliably identified and effectively cleaned. DBScope currently enables artifact screening in an iterative process, where the user alternates between visualization and filtering/cleaning steps. However, this approach is time-consuming and ineffective when the entire frequency spectrum is affected. In this respect, we are invested in the development of algorithms that not only automatically detect the source of these artifacts, but also clean them accordingly.

One of our major goals was to create the conditions for the DBS community to adopt and easily contribute to the enhancement of this toolbox, in response to advancements and discoveries in DBS research. For this reason, DBScope is “open source” and is accessible through an online repository. We firmly believe that these types of initiatives are key in fostering the emergence of novel methods for clinical integration. Although we are currently working on updates, we encourage the clinical and research communities to adapt the available tools and to share their insights, becoming part of this joint effort to improve the DBS therapy.

## Conclusion

DBScope is an open-source computational toolbox to import, visualize and analyze files from the Percept PC device (Medtronic, BrainSense Technology). The toolbox can be used programmatically or through an interface for users without programming experience. This way, it can be directly integrated into the clinical and research practices, whilst remaining adaptable to one’s research questions. Its functionalities are up to date with current literature standards for the evaluation of LFP in DBS. Moreover, future updates are in store. Overall, DBScope is a versatile tool focused on the widespread improvement of data-driven DBS therapies, by expanding the accessibility of the data and by promoting new forms of analyzing the complexity of DBS data. The software, as well as a detailed user guide and a sample test dataset, can be downloaded at https://github.com/NCN-Lab/DBScope.

## Supporting information

SuppInformation

## Data Availability

All data produced are available online at https://github.com/NCN-Lab/DBScope

## Acknowledgments

We would like to express our sincere gratitude to Eng. Gaetano Leogrande from Medtronic, for his invaluable insight and suggestions to the development of the presented computational toolbox. Figures 1 and 3 were created with BioRender.com.

## Author contributions

A.O., E.C. and P.A. conceived and developed the presented toolbox. A.O., E.C. and P.A. designed and analysed the data in the case studies. A.O., E.C. and P.A. conceived and wrote the manuscript draft. All co-authors critically reviewed the manuscript. P.A. supervised the research.

## References

1. Krauss JK, Lipsman N, Aziz T, et al. Technology of deep brain stimulation: current status and future directions. Nature Reviews Neurology 2020 17:2 2020;17(2):75–87.

2. Miocinovic S, Somayajula S, Chitnis S, Vitek JL. History, Applications, and Mechanisms of Deep Brain Stimulation. JAMA Neurology 2013;70(2):163.

3. Okun MS. Deep-Brain Stimulation for Parkinson’s Disease. https://doiorg/101056/NEJMct1208070 2012;367(16):p1529-1538.

4. He S, Baig F, Mostofi A, et al. Closed-Loop Deep Brain Stimulation for Essential Tremor Based on Thalamic Local Field Potentials. Movement Disorders 2021;36(4):863–873.

5. Johnson MD, Miocinovic S, McIntyre CC, Vitek JL. Mechanisms and targets of deep brain stimulation in movement disorders. Neurotherapeutics 2008;5(2):294–308.

6. Chiken S, Nambu A. Mechanism of Deep Brain Stimulation:Inhibition, Excitation, or Disruption? The Neuroscientist 2016;22(3):313–322.

7. Parastarfeizabadi M, Kouzani AZ. Advances in closed-loop deep brain stimulation devices. Journal of NeuroEngineering and Rehabilitation 2017;14(1).

8. Moldovan AS, Hartmann CJ, Trenado C, et al. Less is more - Pulse width dependent therapeutic window in deep brain stimulation for essential tremor. Brain Stimul 2018;11(5):1132–1139.

9. Dayal V, Limousin P, Foltynie T. Subthalamic Nucleus Deep Brain Stimulation in Parkinson’s Disease: The Effect of Varying Stimulation Parameters. Journal of Parkinson’s Disease 2017;7:235–245.

10. Goyal A, Goetz S, Stanslaski S, et al. The development of an implantable deep brain stimulation device with simultaneous chronic electrophysiological recording and stimulation in humans. Biosensors and Bioelectronics 2021;176:112888.

11. Feldmann LK, Lofredi R, Neumann WJ, et al. Toward therapeutic electrophysiology: beta-band suppression as a biomarker in chronic local field potential recordings. npj Parkinson’s Disease 2022;8(1).

12. Neumann WJ, Gilron R, Little S, Tinkhauser G. Adaptive Deep Brain Stimulation: From Experimental Evidence Toward Practical Implementation. Movement Disorders 2023.

13. Thenaisie Y, Palmisano C, Canessa A, et al. Towards adaptive deep brain stimulation: clinical and technical notes on a novel commercial device for chronic brain sensing. Journal of Neural Engineering 2021;18(4):042002.

14. Stam MJ, van Wijk BCM, Sharma P, et al. A comparison of methods to suppress electrocardiographic artifacts in local field potential recordings. Clinical Neurophysiology 2023;146:147–161.

15. Cagle JN, Johnson KA, Almeida L, et al. Brain Recording Analysis and Visualization Online (BRAVO): An open-source visualization tool for deep brain stimulation data. Brain Stimulation 2023.

16. Hammer LH, Kochanski RB, Starr PA, Little S. Artifact Characterization and a Multipurpose Template-Based Offline Removal Solution for a Sensing-Enabled Deep Brain Stimulation Device. Stereotactic and Functional Neurosurgery 2022;100(3):168–183.

17. Chen Y, Gong C, Tian Y, et al. Neuromodulation effects of deep brain stimulation on beta rhythm: A longitudinal local field potential study. Brain Stimulation 2020;13(6):1784–1792.

18. Gilron Re, Little S, Perrone R, et al. Long-term wireless streaming of neural recordings for circuit discovery and adaptive stimulation in individuals with Parkinson’s disease. Nature Biotechnology 2021;39(9):1078–1085.

19. Guidetti M, Marceglia S, Loh A, et al. Clinical perspectives of adaptive deep brain stimulation. Brain Stimulation: Elsevier Inc.; 2021. p. 1238–1247.

20. Neumann WJ, Turner RS, Blankertz B, Mitchell T, Kühn AA, Richardson RM. Toward Electrophysiology-Based Intelligent Adaptive Deep Brain Stimulation for Movement Disorders. Neurotherapeutics 2019 16:1 2019;16(1):105–118.

21. Merk T, Peterson V, Köhler R, Haufe S, Richardson RM, Neumann WJ. Machine learning based brain signal decoding for intelligent adaptive deep brain stimulation. Experimental Neurology 2022;351:113993–113993.

