## Supplementary material for "DBScope: a versatile computational toolbox for the visualization and analysis of sensing data from Deep Brain Stimulation": SuppInformation

```matlab
%% DBScope example script
% Initialize object
obj = NCNPERCEPT_BATCH;
[ status, text ] = obj.open_batch_files();
% Exemplify with one file
patient = 1; file = 1;
% Disp Lead Information
text = obj.ncnpercept_patient{patient}{file}.getLeadConfig();
% Chronic - Timeline
obj.ncnpercept_patient{patient}{file}.chronic_obj.plotLFPTrendLogs();
% Chronic - Circadian Distribution
obj.ncnpercept_patient{patient}{file}.chronic_obj.plotCircadian();
% Online Streaming - ECG Artifact Cleaning
obj.ncnpercept_patient{patient}{file}.streaming_obj.cleanECG(250);
% Online Streaming - Data Summary
obj.ncnpercept_patient{patient}{file}.streaming_obj.plotSummaryStreaming(1);
```

**Figure S1. DBScope *script example.*** The initial steps involve initializing the mother object, NCNPERCEPT_BATCH, followed by the file(s) loading. Subsequently, one file is analyzed considering three different streams of operations: Calibration System, for the display of the lead information; Chronic Sensing, for the visualization of the Timeline and circadian distribution; and Online Streaming Sensing, for the application of the ECG artifact cleaning algorithm and visualization of the data summary.

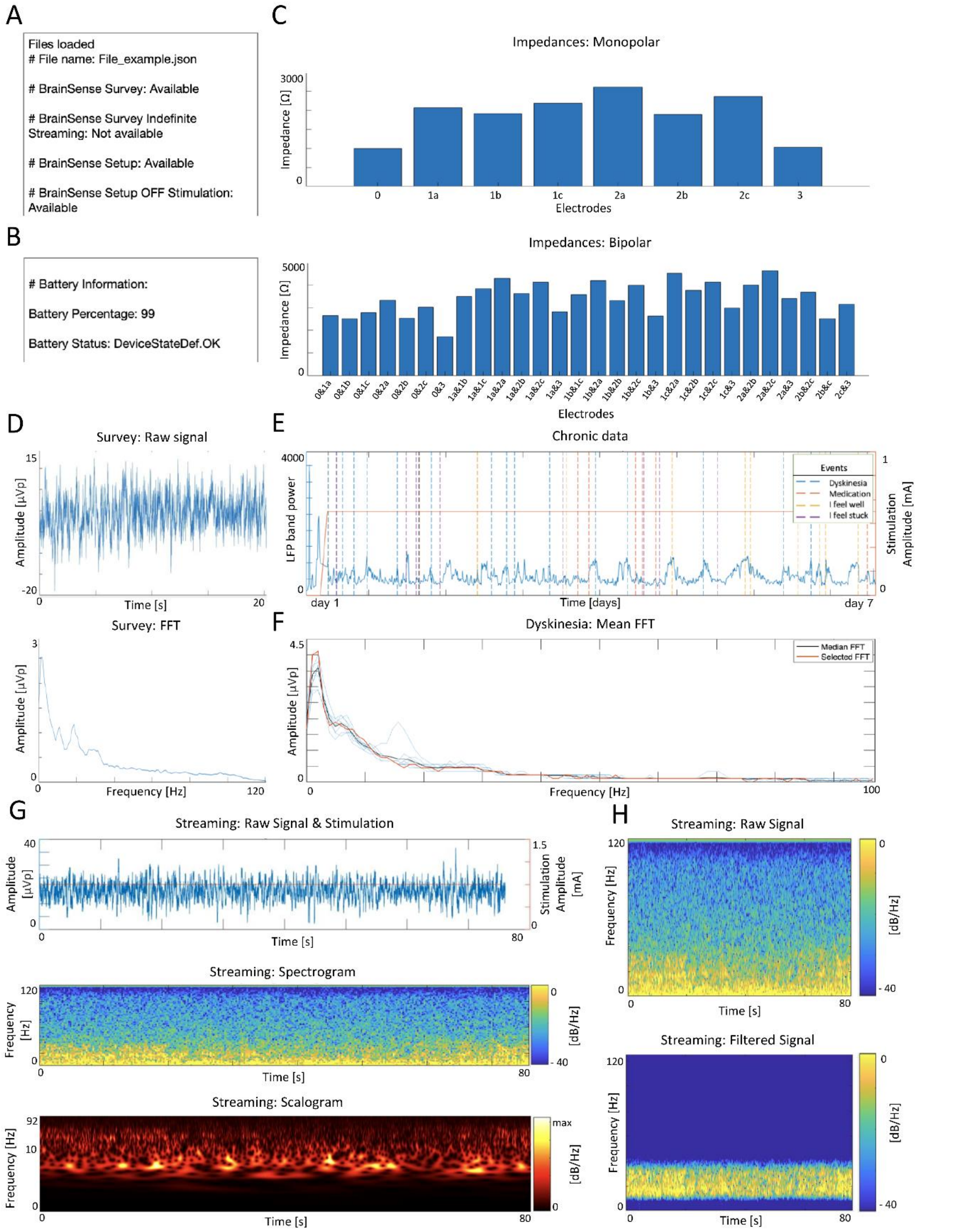

**Figure S2: Overview of the toolbox outcomes. (A)** Snapshot of the text window displayed once a file is loaded.**(B)** Snapshot of the battery information. **(C)** Impedance information (monopolar and bipolar) for one hemisphere, accessible in the Calibration section. **(D)** Raw signal and Fast Fourier Transform (FFT) of a Survey recording. **(E)** Chronic data of one hemisphere, with the events marked by the patient. **(F)** FFT of 'Medication episodes marked in **(E)**. **(G)** Data summary, containing the raw signal, the stimulation amplitude, the spectrogram, and the scalogram obtained with a wavelet method of a Online Streaming recording. **(H)** Spectrograms of the recording of (G) before and after a bandpass filter.

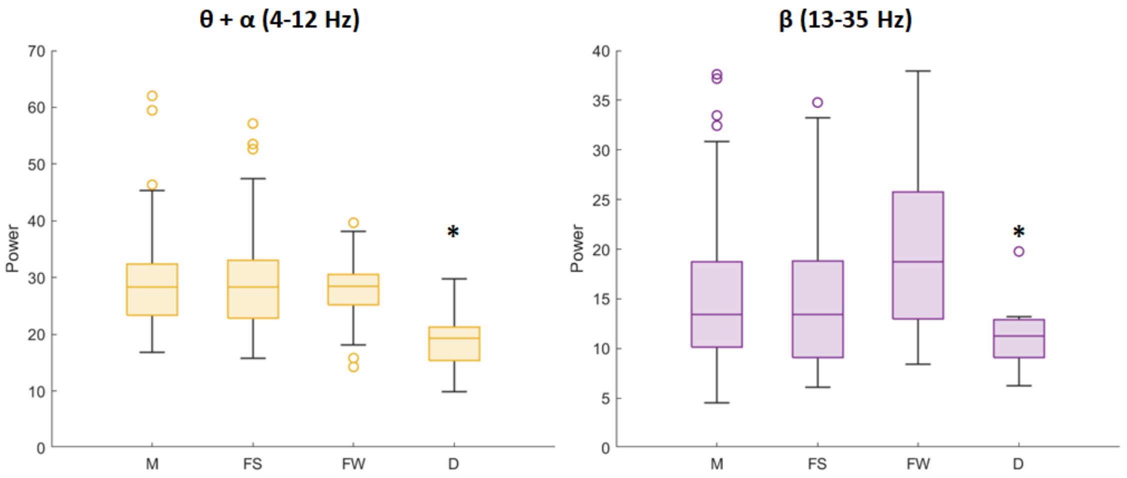

**Figure S3 - Power in alpha-theta and beta frequency bands of four different events.** Each graph has the minimum, 25th percentile, median, 75th, and maximum values for the four types of events: M – Medication; FS – Feeling stuck; FW – Feeling well; D – Dyskinesias. The asterisk marks the instances where dyskinesias are statistically different from at least one other event (ANOVA, $p<0.05$).

### USER GUIDE

#### DBScope

Visualization and Analysis of DBS data from Percept PC

Toolbox for import, preprocessing, visualization and analysis of Deep Brain Stimulation sensing recordings extracted from Medtronic's Percept PC

Andreia M. Oliveira, Eduardo Carvalho, Beatriz Barros & Paulo Aguiar
Faculty of Engineering of University of Porto (FEUP)
Neuroengineering and Computational Neuroscience (NCN) Lab,
*I3S – Instituto de Investigação e Inovação em Saúde*
Porto, Portugal
[]
2023

### Installation

To install DBScope, you need to have MATLAB installed. The toolbox operates better in versions MATLAB_R2021b and upwards. If your version is older than this one, the code can be incompatible at times. MATLAB is a paid application. To consult the available offers, please refer to https://www.mathworks.com/products/matlab.html .

Download the DBScope package from https://github.com/NCN-Lab/DBScope

Once you have MATLAB installed, open the programming environment. Open the directory in the folder containing the DBScope package:

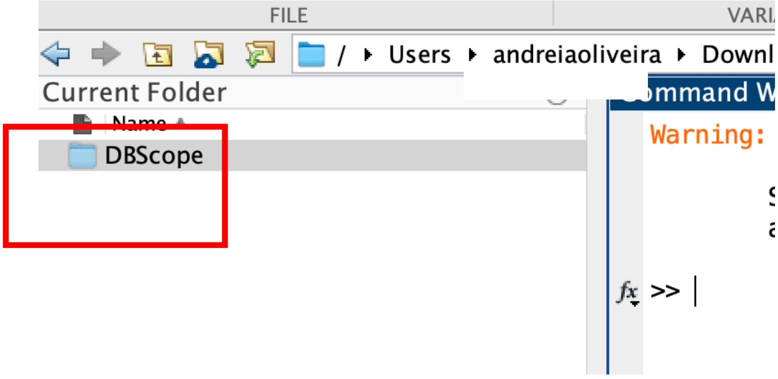

Then, add the 'DBScope' to path, by selecting the option 'Add to Path > Select Folders and Subfolders':

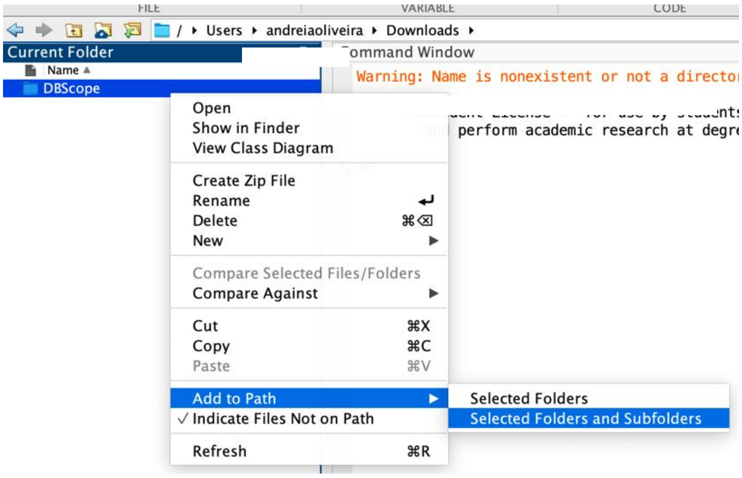

From this moment on, the toolbox is operational. You can access the tools directly via command window:

```
obj = NCNPERCEPT_BATCH;
data_type = obj.aux_open_batch();
switch data_type
    case 'folder'
        [ status, text ] = obj.open_batch_folder();
    case 'files'
        [ status, text ] = obj.open_batch_files();
    case 'workspace'
        [ status, text] = obj.open_mat_files();
end
```

Or through the User Interface. To open the User Interface, run the command:

**`>> DBScope`**

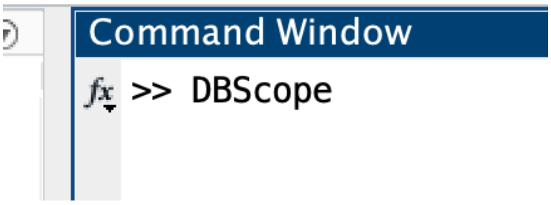

#### List of equipment/material/hardware

The input data of the toolbox are the *.json* files directly extracted from the clinical console of the Percept PC. No additional equipment is needed. Some analyses are computational demanding, and may run slower.

### DBScope Package

The DBScope toolbox was built for the import, preprocessing, visualization and analysis of Deep Brain Stimulation sensing recordings extracted from Medtronic's IPG - Percept PC. The files contain several types of recordings corresponding to the sensing modes available. The Percept PC has four main sensing modes: survey and setup, for in-clinic parameterization of the leads for sensing and stimulation; streaming, for in-clinic recordings; and timeline with events, for out-of-clinic recordings, containing snapshots of events logged by the patient.

Besides the graphical user interface that allows to easily access the information available in the data structure, the tools can be accessed programmatically (script and command window), bypassing the user interface. A step-by-step guide for these functionalities will be described below, with examples.

#### 1) Main graphical user interface

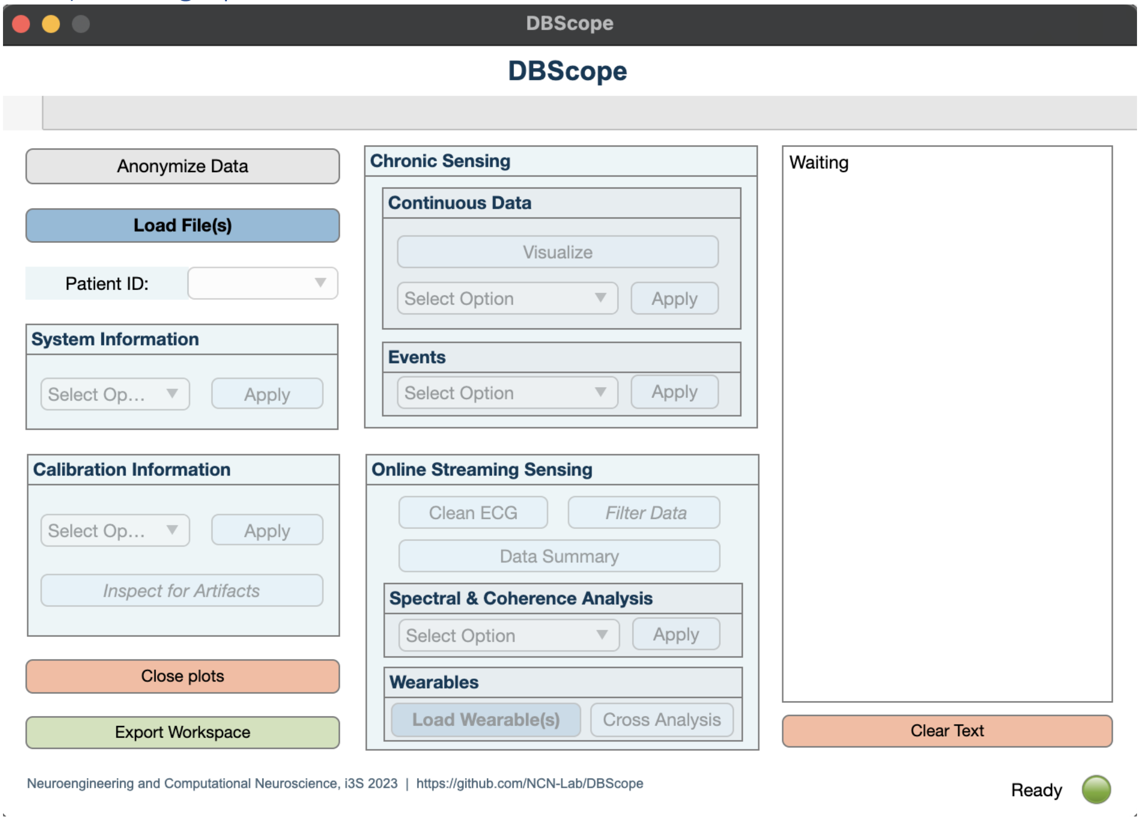

**Figure 1.** ***DBScope:*** Toolbox for import, preprocessing, visualization and analysis of Deep Brain Stimulation sensing recordings extracted from Medtronic's Percept PC – main window.

The toolbox can operate either on single file or multiple files (multi-selected or by uploading a folder). It also possible to upload previously recorded workspaces in *.mat* format.

A note on the visualization window: as each visualization window is independent, is it possible to open and operate several visualization windows simultaneously.

To load the file(s) click on the button '**Load File(s)**' and select the process and files to upload.

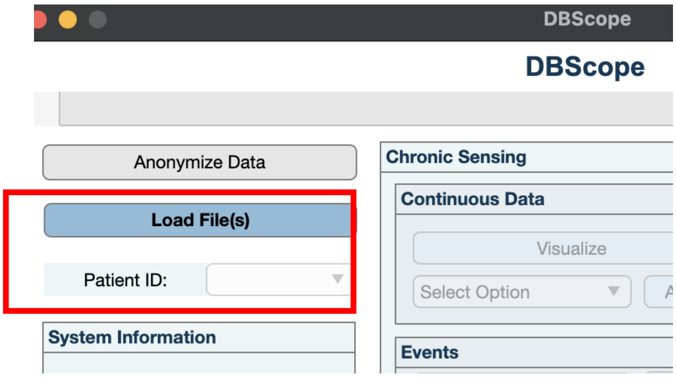

Once the file(s) is(are) uploaded, a message will appear in the text box with the information of which sensing recordings modes are available:

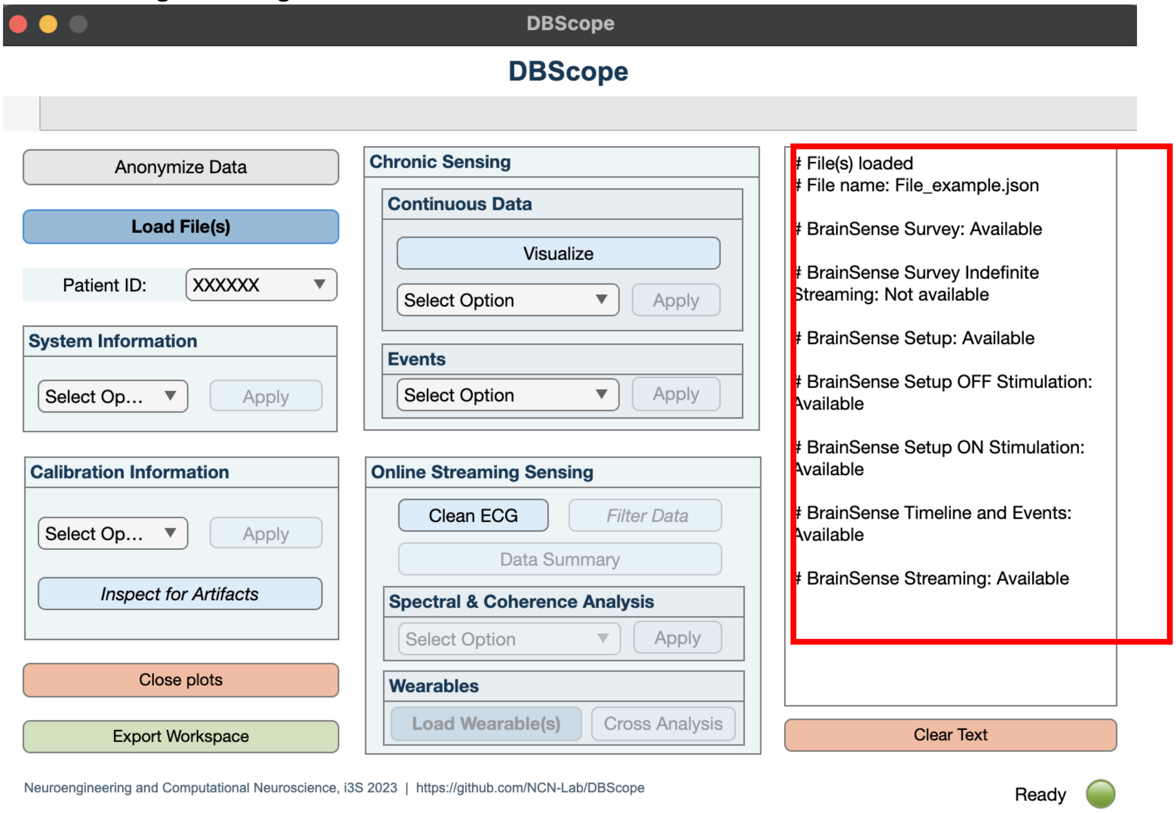

Depending on the sensing modes in the files, the following features are available: System Information, Calibration Information, Chronic Sensing and Online Streaming Sensing.

#### 2) System Information

Here, the user can access system, patient, and hardware information. The information will be displayed in the text window on the right.

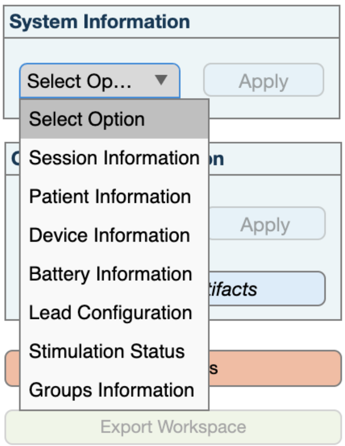

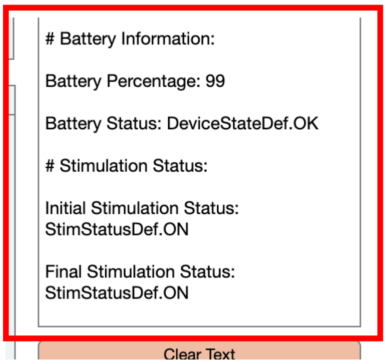

#### 3) Calibration Information

In this section, the user can access information about calibration. This includes the impedance tests performed in clinic, the presence of artifacts, and plots of the recordings from the recording modes Survey, Setup and Indefinite Streaming (if available in file(s)). The information about the presence of artifacts detected by the IPG will be made available in the text window.

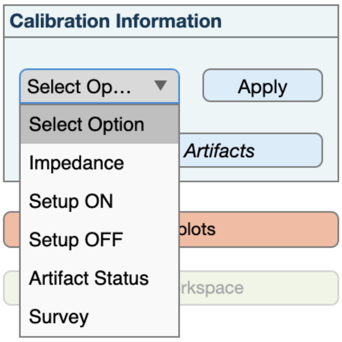

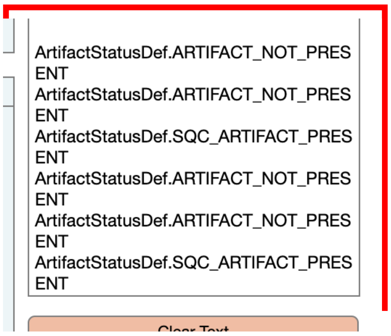

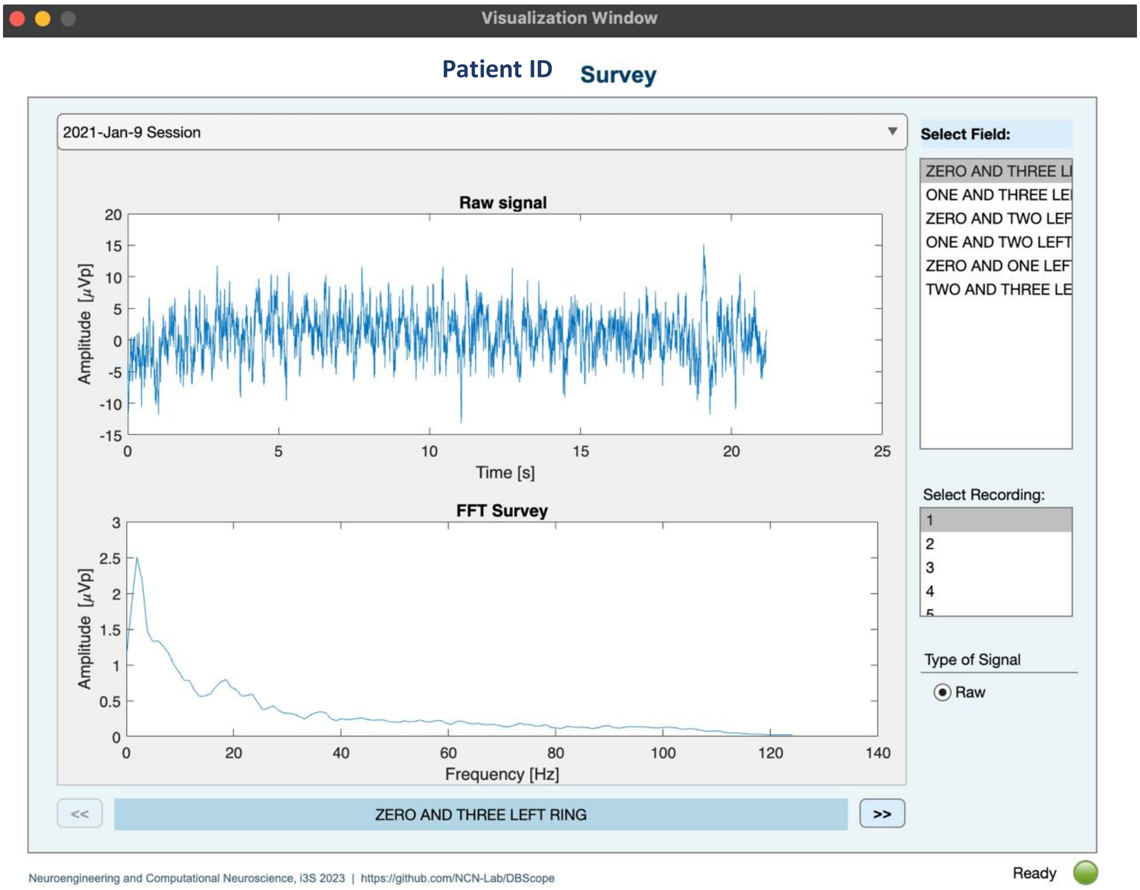

[Example of Survey recording visualization.]

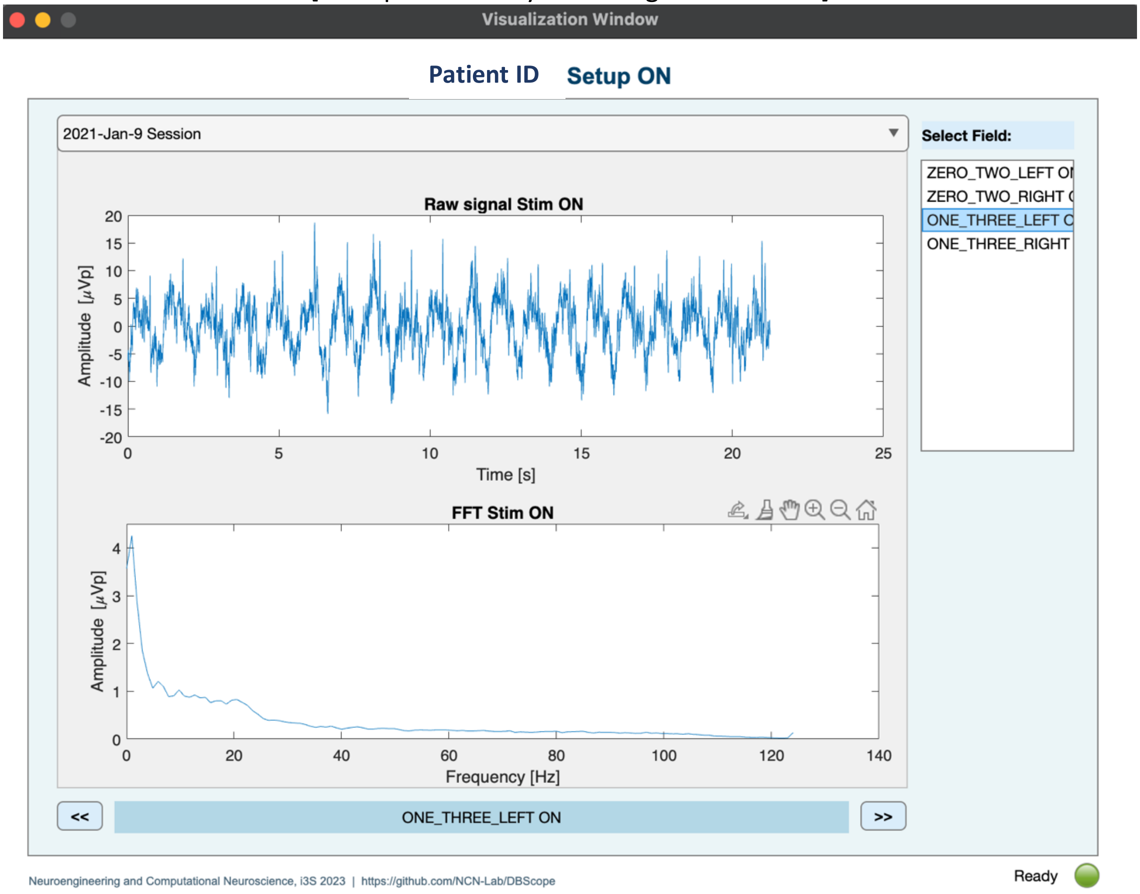

[Example of Setup Stimulation ON recording visualization.]

#### Impedance

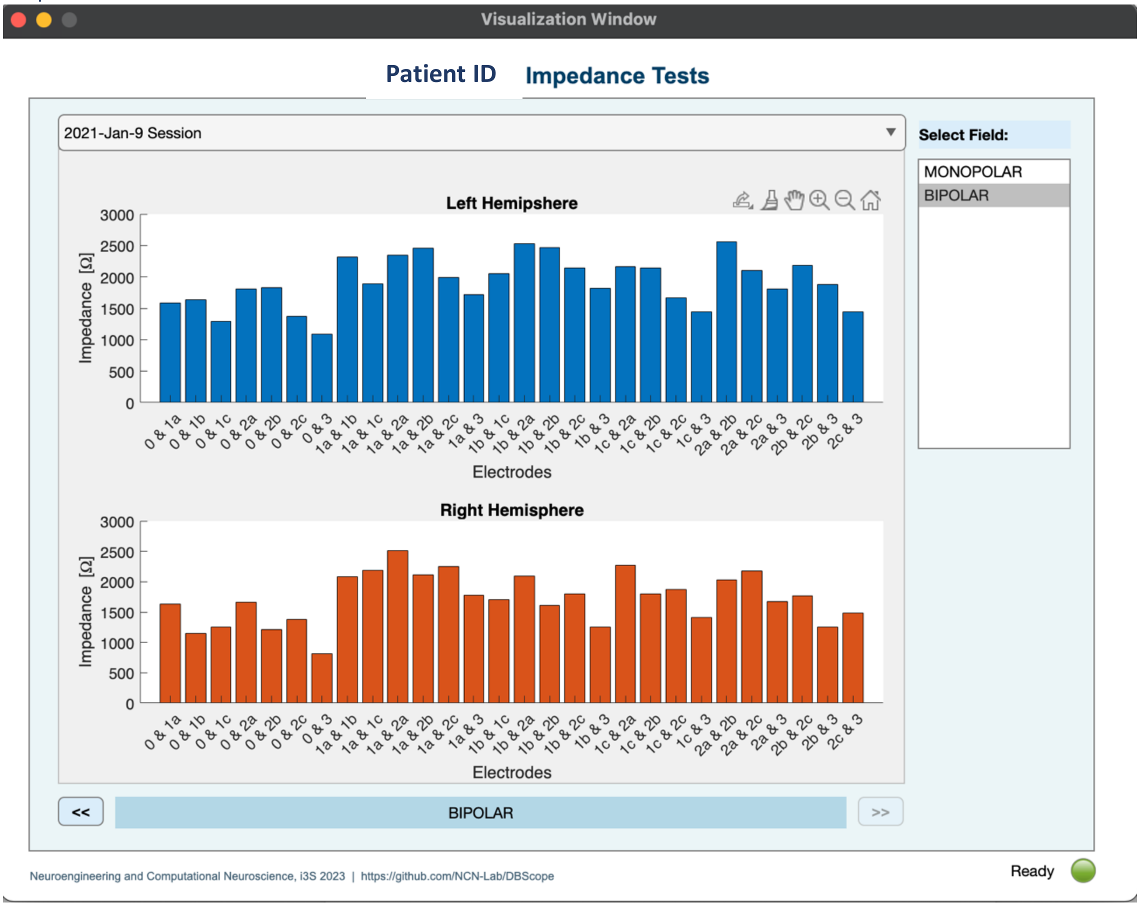

[Example of Impedance visualization.]

#### Inspect for Artifacts

A secondary User Interface is available at **'Inspect for Artifacts**'. This feature allows the user to visualize the survey signals both in the time and frequency domains, and to design custom filters, that may help in the manual inspection for artifacts.

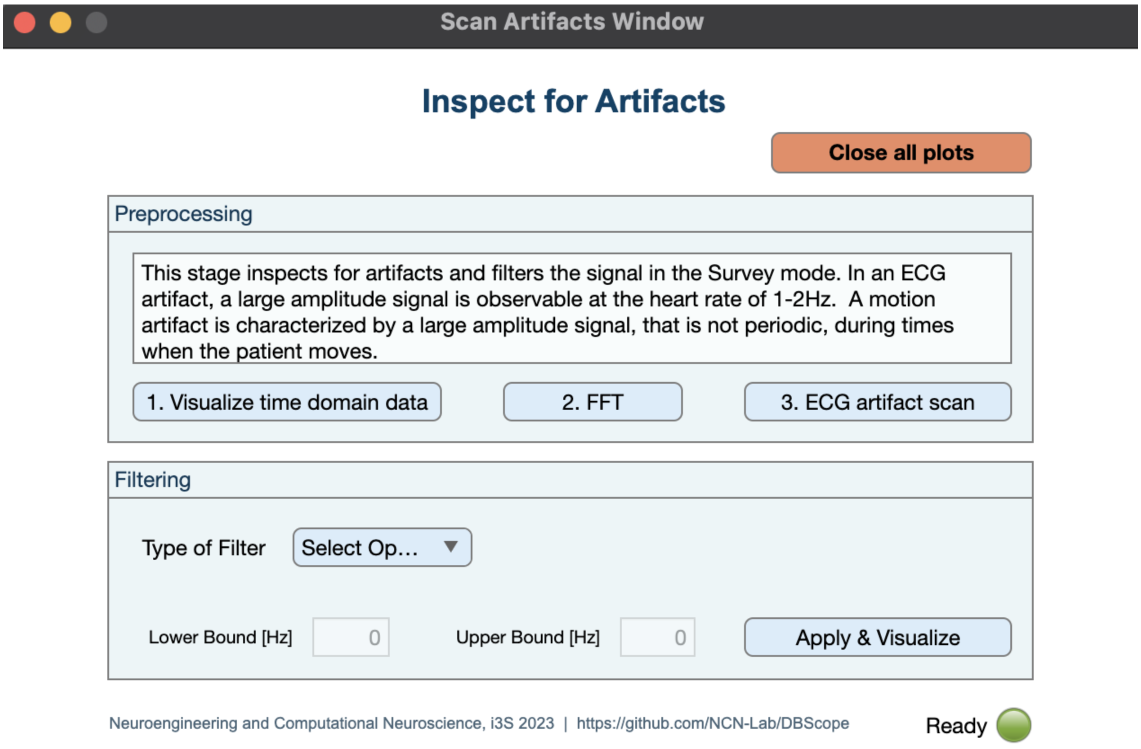

Figure 2: ***DBScope:*** Toolbox for the inspection of artifacts in survey sensing recordings – secondary window.

#### 4) Chronic Sensing

In the section for chronic sensing recordings, one can access two main types of data: timeline and events snapshots.

##### Timeline

Timeline recordings can be plotted at the button '**Visualize**':

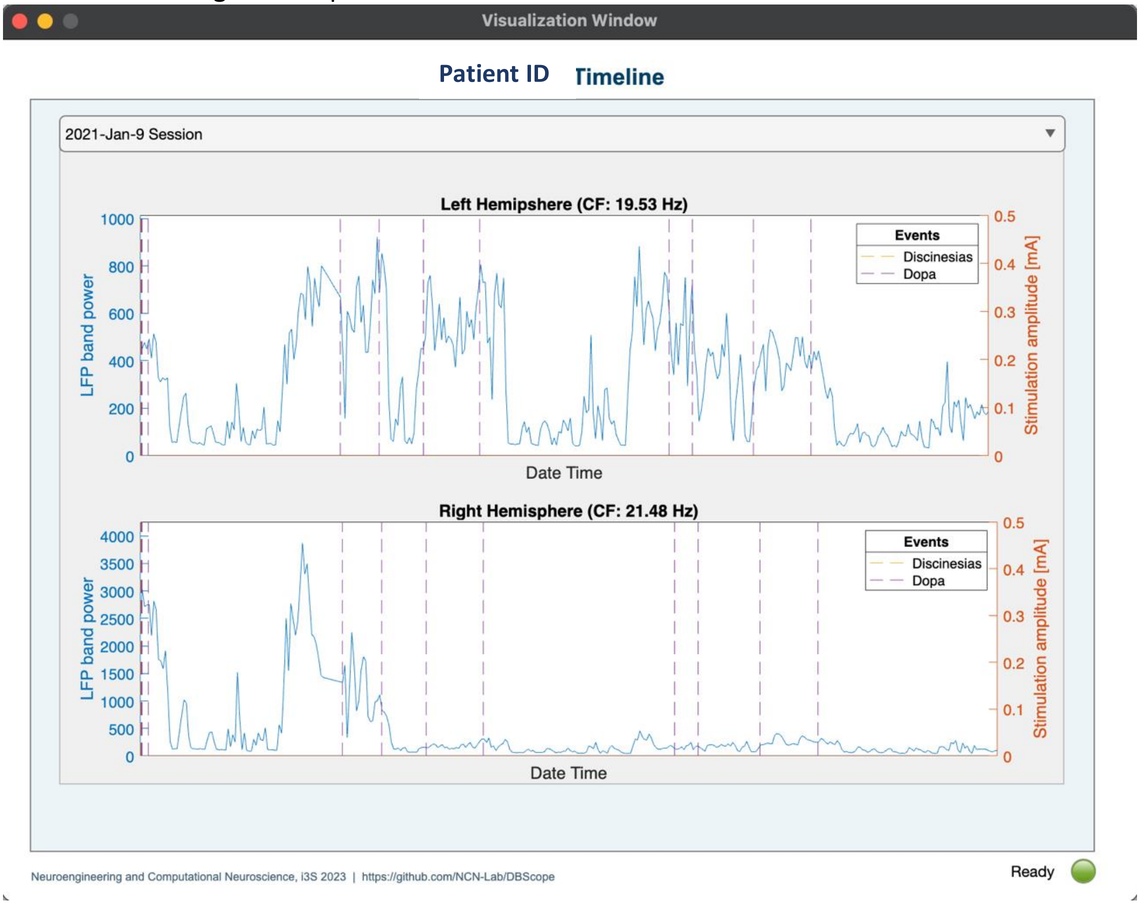

The user can also perform further analysis on chronic data:

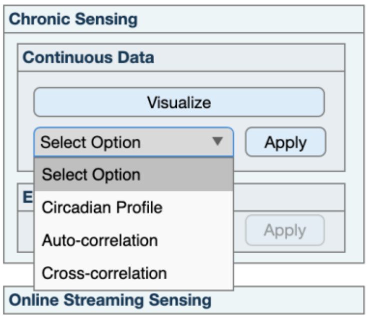

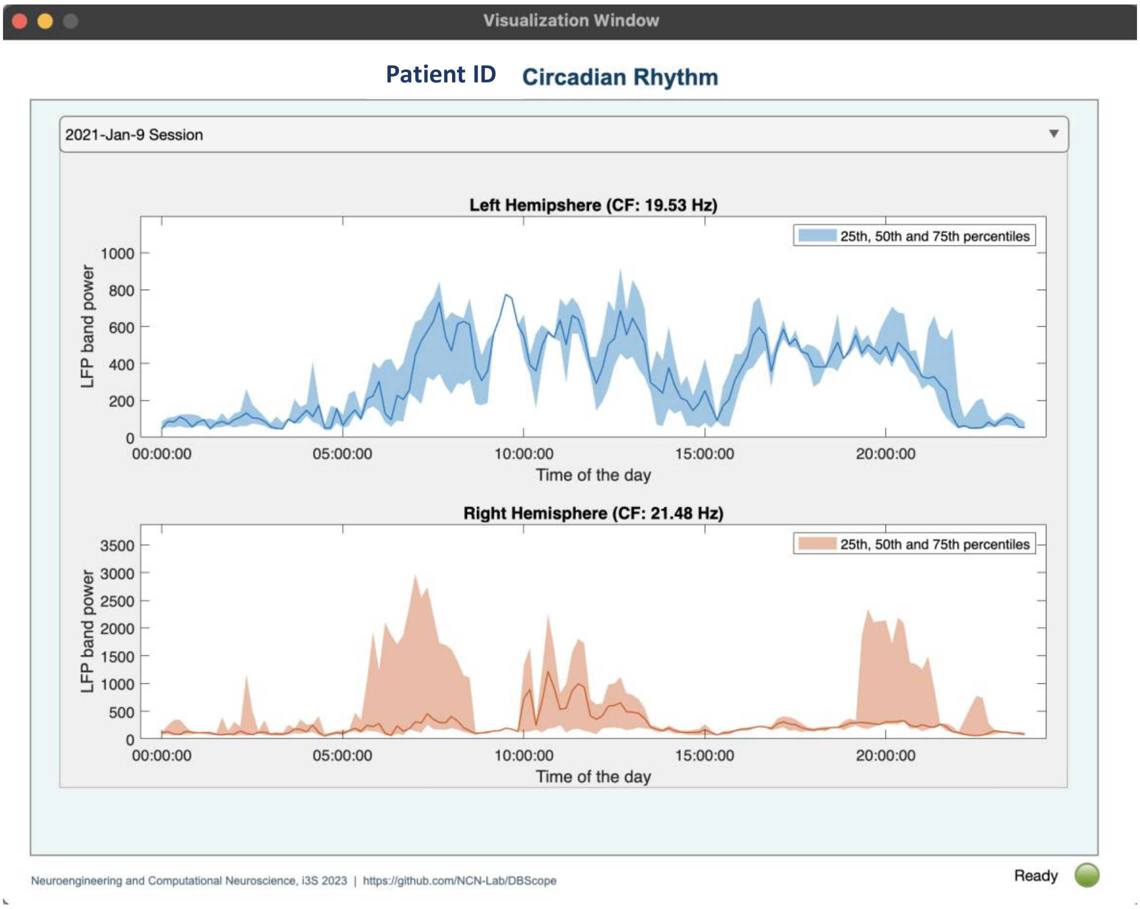

[Example of Circadian Profile visualization.]

#### Events

A second type of chronic recordings are the events logged by the patients. For the events, the following analyses are available: the Fast Fourier Transform (FFT) per event, Event-Triggered average, and the information of their temporal dynamics.

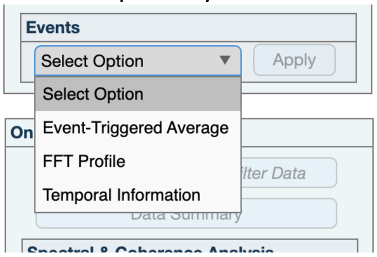

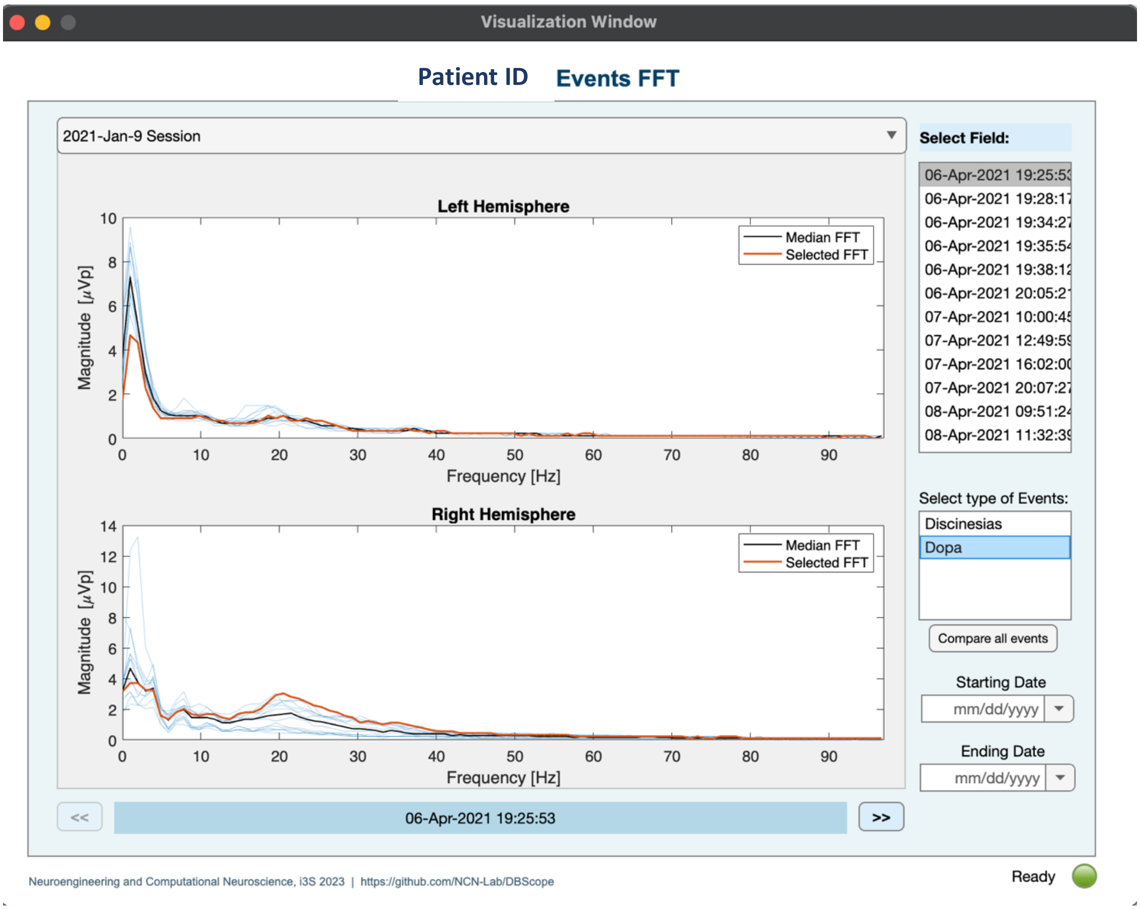

[Example of Events FFT visualization.]

Depending on the characteristics of the recordings, it is possible, in the visualization window, to change files, define a temporal window, select the event type, and individual recordings.

#### 5) Online Streaming sensing

For the online (in-clinic) streaming recordings, a first step is required: cleaning potential ECG artifacts (button 'Clean ECG'). From here onwards, two types of data will be available for plotting and analysis: raw or the ECG-cleaned signals.

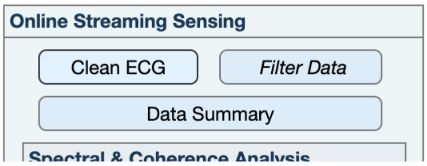

After '**Clean ECG'**, the user can 1) filter the data; 2) plot a **'Data Summary ',** which shows, in the same visualization window, the plot of raw signal, its spectrogram and corresponding Wavelet Transform; and 3) make use of different Spectral and Coherence Analysis tools.

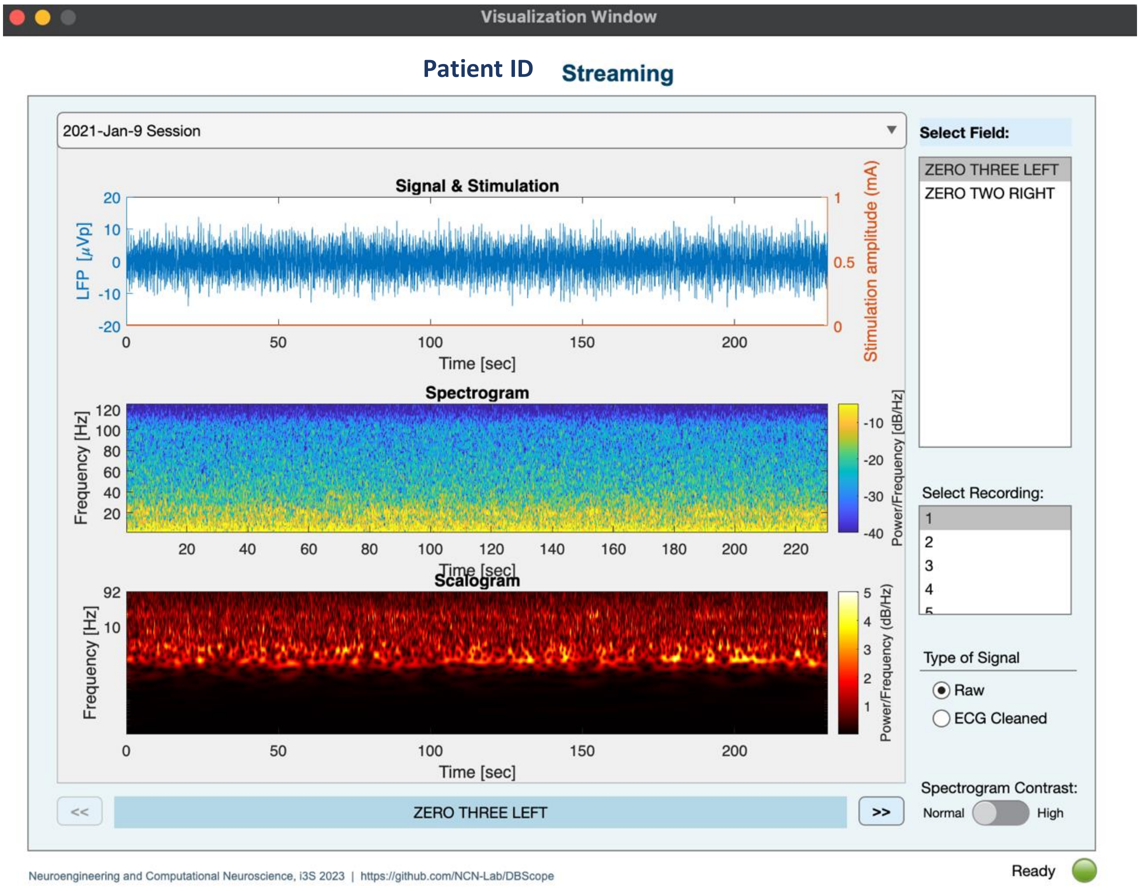

[Example of Data Summary visualization.]

Depending on the characteristics of the recordings, it is possible, in the visualization window, to change files, electrode pairs and recordings. For example, in the visualization above the file is 'File_example', the recording is the '1', and the field (electrode pair) is 'ZERO TWO RIGHT'.

#### Filter data

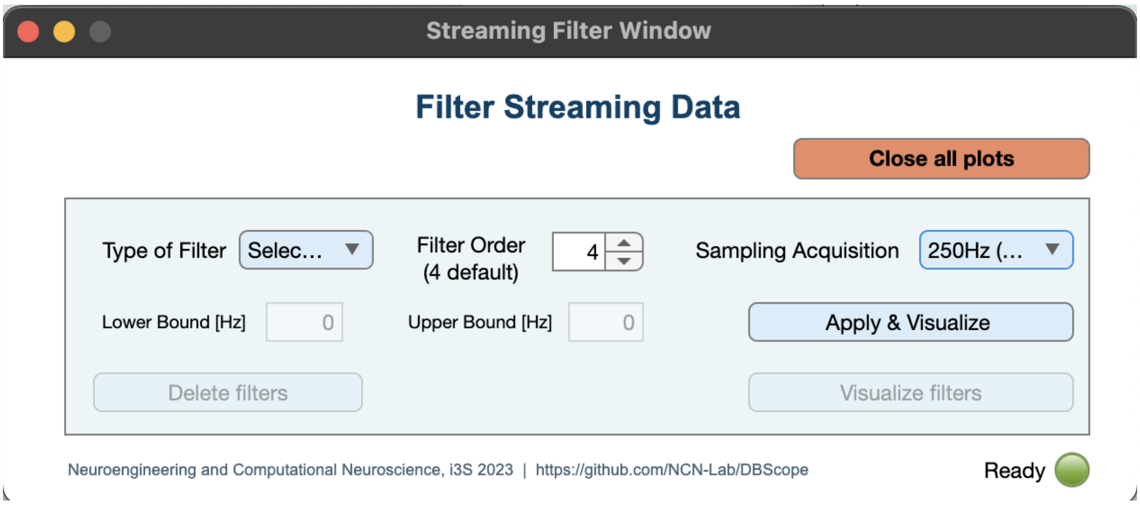

**Figure 3:** ***DBScope:*** Toolbox for filter design and application in streaming sensing recordings – secondary window.

In this secondary window, the user can define the type of filter (low pass, high pass, stopband and bandpass), and its boundaries (below the Nyquist frequency of 125Hz). It is possible to test an unlimited number of filters, as well as delete previous filtering iterations. The output of this process will then be available in the main GUI.

#### Spectral and Coherence Analysis

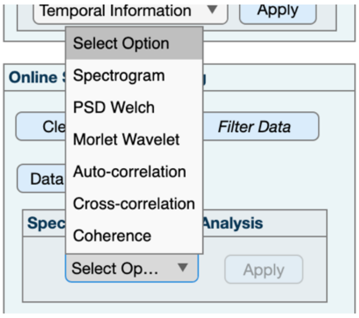

The set of tools available in the Spectral and Coherence Analysis tools can be applied to either the raw, the ECG-cleaned or the filtered data.

#### 6) External Wearables

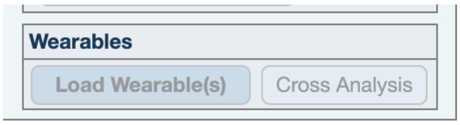

The set of tools available in the Wearables section allows for the load and cross visualization of the wearable data, simultaneously to the visualization of the corresponding streaming recordings.

##### Loading of wearable data

The data must be in a .CSV file named:

WearableID _PatientID_Task_TitrationInfo_SamplingFreq_StartDatetime('yyyy-MM-ddTHH-mm-ss').csv

Example: 'DorsalLeft_12345_OpenClose_RightTitration_100_2023-05-19T08-58-05.csv'

The first column of the file must contain the datetime in a ".net" format:

Number of "clock ticks" since 1-Jan-0001 00:00:00 UTC, representing a Microsoft® .NET timestamp where each clock tick is 100 ns.

Besides the datetime column, the file can contain as many columns as you wish. Each column will be treated as a separate signal. For instance, you can have the 3-axis acceleration and 3-axis magnetometer values within the same file. Note that DBScope will use the name of the columns to identify the signals within its data structure.

When in the presence of metrics with different sampling frequencies, choose the higher sampling frequency and fill the other values with NaN:

| 1 | time | total_acc | total_acc_power |
|---|---|---|---|
| 2 | 638200896040100000 | NaN | NaN |
| 3 | 638200896040200000 | NaN | NaN |
| 4 | 638200896040300000 | 1.00950273821584 | NaN |
| 5 | 638200896040400000 | 1.00796206052089 | NaN |
| 6 | 638200896040500000 | 1.01701394986963 | NaN |
| 7 | 638200896040600000 | 1.00377437452695 | NaN |
| 8 | 638200896040700000 | 1.00599844258228 | NaN |
| 9 | 638200896040800000 | 1.01399387544742 | NaN |
| 10 | 638200896040900000 | 1.01965266999158 | NaN |

##### Cross Analysis

When performing cross-signal analysis, DBScope will automatically select the wearable recordings that overlap the currently selected *Streaming* recording. The signals are aligned and limited by the *Streaming* recording datetime window, so that wearable recordings that start before or end after the *Streaming* recording are only partially shown.

**NOTE**: Make sure that the neurostimulator and wearables are synchronized.

#### 7) Export Workspace & Close App

The user can also save his analysis with the '**Export Workplace**' button. This step allows to save the current operational workplace in a *.mat* file.

Finally, when closing the app, the following prompt window will appear:

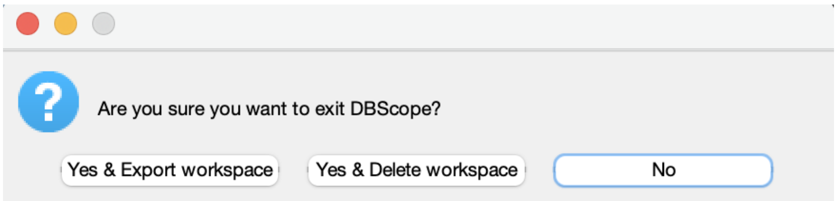

Here, the user can select to leave the app - and export or delete the current workspace -, or to continue in the app.

### Contact Us

For questions / comments, please send us an email to:

, or
